# A machine learning model to explore individual risk factors for tuberculosis treatment non-adherence in Mukono district

**DOI:** 10.1101/2022.12.01.22283003

**Authors:** Haron W. Gichuhi, Mark Magumba, Manish Kumar, Roy William Mayega

## Abstract

Despite the availability and implementation of well-known efficacious interventions for Tuberculosis (TB) prevention and treatment by the Ministry of Health, Uganda (MoH), treatment non-adherence persists. Moreover, identifying a specific patient at risk of non-adherence is still a challenge. Thus, this study set out to utilize machine learning modeling to explore individual risk factors predictive of tuberculosis treatment non-adherence in the Mukono district.

This was a retrospective study based on a record review of 838 TB patients enrolled in six health facilities (3 government, 3 private-not-for-profit) in the Mukono district. We developed five machine learning algorithms (Logistic regression (LR), artificial neural networks (ANN), support vector machines (SVM), Random Forest (RF), and AdaBoost) to explore the individual risk factors for tuberculosis treatment non-adherence. Also, we evaluated their accuracy by computing the positive predictive value (PPV) and negative predictive value (NPV) through the aid of a confusion matrix.

Of the five developed and evaluated models, SVM performed the best with an accuracy of 91.28 % compared to RF (89.97%), LR (88.30%), ANN (88.30%), and AdaBoost (91.05%) respectively. Individual risk factors predictive of non-adherence included; TB type, GeneXpert results, sub-country, ART status, contacts below 5 years, health facility ownership, sputum test results at 2 months, treatment supporter, CPT Dapson status, risk group, patient age, gender, middle and upper arm circumference, referral, positive sputum test at 5 months and 6 months.

This study shows that classification machine learning techniques can identify patient factors predictive of treatment non-adherence and accurately differentiate between adherent and non-adherent patients. Thus, TB program management should consider adopting the machine learning techniques evaluated in this study as a screening tool for both identity and target-suited interventions for these patients.

## Introduction

Tuberculosis (TB), a curable and preventable infectious disease remains a public health challenge leading to serious economic and social consequences (1). In 2020 alone, there were 1.3 million estimated deaths of tuberculosis patients who were HIV-negative, up from 1.2 million in 2019 globally, according to the World Health Organization’s report (1). In Africa, approximately 1.6 million new cases and over 600,000 deaths occur each year (1).

Uganda, like other African countries, is still facing tuberculosis challenges. For instance, in 2019 alone, 88,000 people were infected with TB and an estimated 15,600 people died (2) in the same year. Yet, only 61% of people with TB symptoms seek appropriate care from health facilities (2). Although TB treatment coverage has greatly improved from 65% (2015/2016) to 76% (2020/2021) in the last 5 years, this is still below the national target of 90% (2).

One of the major tuberculosis management challenges is the TB treatment non-adherence and the consequent difficulty in identifying these non-adherent patients (2–4). Treatment non-adherence, as defined by the World Health Organization (WHO) is the extent to which a person does not conform to the TB treatment regime as recommended by the health care provider (5). Treatment non-adherence adversely affects the treatment success rate of any disease condition.

For instance, in Uganda, the TB treatment adherence rate is estimated to be 78% while the treatment success rate stands at 72% (2). Thus, the persistent occurrence of TB treatment non-adherence, currently estimated at 22%, despite the availability and implementation of well-known efficacious interventions for TB prevention and treatment by the Ministry of Health, Uganda (MoH) (6), implies the possible existence of factors intrinsic to the patients or the treatment strategies amongst others that could be hindering adherence.

Globally, the WHO guidelines recommend active pulmonary TB patients strictly take and complete a drug regimen usually administered in two phases (intensive and continuation phase). Further, the patients must achieve a treatment adherence level greater than 90% in 6-9 months (7) (8). Recently, shorter drug regimens for four months have been approved (9). However, this TB treatment is associated with serious side effects with the commonest being nausea (74%), diarrhea (46%), hearing loss (16%), and psychosis (12%). As a result, some patients do not complete their medications (10–13). Failure to which has been found to reduce treatment success, increase the risk of the patients developing drug-resistant strains, burden the health systems (14), increase family financial hardships (14,15), and spread TB in the community. This in turn increases TB morbidity and mortality (7).

The Uganda National Tuberculosis and Leprosy Program (NTLP) has made significant strides in TB prevention and treatment. For instance, to promote TB treatment adherence, it has adopted and implemented the WHO-recommended strategies namely directly observed treatment therapy (DOT) and video observed therapy (VOT) country-wide. Although NTLP has adopted the WHO treatment adherence strategies, there is no specific way to measure how intensive health providers’ support and supervision should be and to which TB patients it should be provided. Currently, a health worker manually reviews the patient’s medical records and intuitively tries to estimate whether the patient will adhere to treatment or not. This method is cumbersome, time-consuming, and prone to human error. Consequently, many patients go unidentified.

Worse still, Uganda is ranked among the top 30 high-burden TB/HIV countries in the world (1). This ranking shows that Uganda may not achieve the world health organization (WHO) END TB strategy targets of treatment coverage of ≥ 90% and a treatment success rate of ≥ 90 % by 2030 (16). Therefore, efforts and interventions targeting people with TB geared towards treatment seeking, uptake, and adherence are much needed.

Notably, such interventions as early diagnosis and effective treatment are key in stopping the spread of TB and improving patient treatment outcomes (17). Effective, free-of-charge tuberculosis cure exists at designated public and private health facilities through the government of Uganda’s initiative, STOP-TB program – aiming to diagnose and initiate patients early on to TB treatment - implemented by the MoH countrywide (18).

However, for effective TB treatment, the patients must achieve a treatment adherence level greater than 90% in 6-9 months (7). Failure to which has been found to reduce treatment success, increase the risk of the patients developing drug-resistant strains, burden the health systems (14), increase family financial hardships (14,15), and spread TB in the community. This in turn increases TB morbidity and mortality (7).

Reasons for TB treatment non-adherence are complex and influenced by the interplay between; the patient, the health workers, and health systems factors that vary by settings (19,20). However, patient factors have been identified to play a central role in treatment non-adherence (21–23).

Evidence from a systematic review of studies conducted between 1966 - 2005 illustrated family characteristics, income, social support, and relationships with health providers as determinants for non-adherence to TB treatment (24). More so, a case-control study conducted in Indonesia found a lack of TB education provided by TB nurses and the use of loose and fixed-dose combinations to be associated with non-adherence to TB treatment (25).

Further, several studies identified other patient-related factors like being male (13,26–28), comorbidity with HIV/Aids and taking ART drugs (27), patient occupation (15,19,29), intensive treatment (4,15,30,31), travel distance to the hospital (3,13,19) greatly affects the adherence level.

Thus, TB management programs have formulated interventions targeted mainly toward the patient. These interventions include; sending daily reminders via phone messages, calling the patients, directly observing the patient whilst taking the treatment (DOT), and, using digital adherence technologies (DATs) such as; ePills dispensers, VOT; for example, 99DOTS (34), and DOT selfie (35). Despite the adoption of these strategies, TB programs still report high treatment non-adherence rates, and increased morbidity and mortality (1,36).

To avert these high treatment non-adherence rates, studies exploring TB treatment non-adherence factors in patients (37–40) have been conducted. These studies employed both traditional statistics and epidemiological approaches utilizing logistic regression and other generalized linear models (15,27,41–43). Whereas these studies identified some factors associated with TB treatment non-adherence, their applicability in identifying an individual tuberculosis patient at risk of treatment non-adherence is limited.

This is mainly because; a) these models are best for making inferences that are aimed at understanding the association between the predictors and the response and not for prediction, and b) Sometimes, the true relationship is more complicated in which case a linear model may not provide an accurate representation of the relationship between the input and output variables. c) the researchers do not split their data into training and testing sets and thus do not evaluate the resultant’ models on raw datasets. d) the traditional statistics are “limited in handling highly dimensional and correlated variables (collinearity assumption)” (44), thus dropping some would be important variables from the resultant models. However, studies conducted in similar contexts have not adequately used machine learning (ML) as a method to generate predictors of treatment non-adherence. Yet, ML models have been proven effective in accurately illustrating the target parameters for implementing stakeholders to ensure adherence to TB treatment (32,33). Therefore, a machine learning approach, incorporating the comprehensive patient and clinical features as captured in the health facility register for modeling, could prove beneficial.

As the name suggests, machine learning (ML) is the creation of computer programs that can learn and therefore improve their performances by gathering more data and experiences (45). However, machine learning promises to improve treatment management, and outcomes by tailoring healthcare to the individual patient.

This promise, of improving treatment management, could be due to the recent advances in computing technologies, low-cost of computer hardware, and the proliferation of electronic medical records systems (EMRs) that has enabled the healthcare industry to generate and store large volumes of patient data, at high speed, and in various formats. In turn, researchers have applied various machine-learning tools and techniques to patient data stored within these EMRs to transform healthcare, personalized medicine, and computer-aided diagnosis (32,46,47).

The reasons for their adoption and utilization include; first, their ability to deal with large, complex, and disparate data, such as that often found within healthcare. Second, the capability to find the associations among different attributes that can affect the outcome of any disease, without subjective preselection, and maximizing data use while minimizing bias. These capabilities prove the suitability of ML techniques for predicting tuberculosis treatment non-adherence.

Notably, the healthcare industry generates highly voluminous, high velocity, and varying formats such as; x-ray images, clinical notes, ultrasound scans, drug prescriptions, admission histories, etc., commonly referred to as the 3V’s (volume, velocity, and variety). These properties make healthcare prime for data mining.

### Use of machine learning in healthcare

Of the chronic diseases, TB, due to its infectious nature, increasing antimicrobial resistance and prevalence in low-and-middle-income (LMIC) countries, for example, India (32,48), Pakistan (49), Morocco (50), Kenya (51), and Uganda (13) has been widely studied. Characteristically, these previous studies gathered demographic and medical histories of a cohort and observed their adherence and outcomes. The researchers then retrospectively applied ML algorithms e.g. random forest, support vector machine (48,49), survival analysis (32,51), and logistic regression (13,50) to determine variables predictive of treatment failure or non-adherence.

For instance, a study carried out in Iran (52) seeking to evaluate and compare different machine learning methods to predict the outcome of the tuberculosis treatment course, used a training dataset (N = 4515) and testing dataset (N=1935) to explore 6 (six) ML algorithms; decision tree (DT), artificial neural network (ANN), logistic regression (LR), radial basis function (RBF), Bayesian networks (BN), and support vector machine (SVM).

To evaluate the algorithms, the investigators computed the prediction accuracy, F-measure, and recall metrics. They found out that decision trees (C4.5) performed the best with model fitness and prediction accuracy of 84.45% and 74.21% respectively. These study findings were similar to studies conducted in Kenya (53) and China (54) that equally reported the decision trees prediction to perform better with an accuracy of 90% and 70.9% respectively. Noteworthy, the study from Kenya concluded that ML techniques have the potential to identify patients at risk for viral failure before their scheduled measurements (53).

In another study (48), where the researchers’ objective was to construct a deep learning model to better target and improve patient care. The researchers trained an ML model in different clinical scenarios to demonstrate its interpretability and adaptability. They utilized 29 features to build four (4) classifiers namely; linear regression, random forest (100 trees and a max depth of 5), support vector machine, and deep neural network. After evaluation, they identified random forests to accurately estimate the weekly missed calls with an accuracy of 72.4%. A related study (55) was carried out in the USA to detect whether a patient will experience an adverse event due to coronary artery disease (CAD) within a 10-year time frame.

In the study, the researchers collected 21,460 patients’ records. Of these, 75% were used for training and 25% for validation. They trained Logistic Regression, Random Forest, Boosted Trees, CART, and Optimal Classification Trees (OCT) classifiers. After evaluation, they found out that random forest was able to identify specific patients with an accuracy of 84.29% closely followed by OCT at 81.54%.

Newer work has explored the utility of advanced machine learning techniques such as support vector machines (SVM), Artificial Neural Networks (ANN), and more for improved classification accuracy. A study by Mian et al., (49) applied SVM in a dataset containing 275 pulmonary tuberculosis symptomatic and confirmed multidrug-resistant (MDR) cases age >=15 with no gender discrimination for feature selection (FS) algorithms to identify and diagnose MDR tuberculosis in Pakistan. The researchers built and evaluated seven (7) classifiers; random forest, k-nearest neighbors, support vector machine, logistic regression, least absolute shrinkage, selection operator (LASSO), artificial neural networks (ANNs), and decision trees. They found out that the two best-performing algorithms were; SVM and RF with an accuracy of 78% and 74% respectively for patients’ classification.

Methodologically, our work was related to other studies predicting outcomes by inferring a model being trained by a set of historical data (32,56,57). Given appropriate assumptions, such techniques allow for valid predictions about the counterfactual outcomes under different settings for determining interventions. However, the ML techniques require exact knowledge of intervention outcomes which should be clearly labeled.

Lastly, this study notes the increasing use of ML for specific customer loan default predictions in the banking sector (58), multilingual tweets classification for disease surveillance (59), and predicting an individual HIV/AIDs patient likely not to adhere to treatment (60). Similarly, this could be replicated in TB management and research.

Further, previous work in this domain has shown that decision trees (DT), random forests (RF), and support vector machines (SVM) perform better in other countries. Unfortunately, based on our research, we did not identify a study utilizing them for TB treatment non-adherence prediction in the Ugandan context. Therefore, our study explored DT, RF, and SVM techniques amongst others.

## Materials and Methods

### Study design and setting

This was a retrospective study of TB patient records conducted in the Mukono district. Within the district, six high-volume (treating > 100 clients) health facilities owned either by the government or private-not-for-profit (PFNP), serving urban, peri-urban, and rural populations were selected. These were; 1) Kyetume Health Centre III (PNFP), 2) Mukono General Hospital (Government), 3) Naggalama Hospital (PNFP), 4) Mukono COU Hospital (PNFP), 5) Kojja Health Centre IV(Government), 6) Nakifuma Health Centre III (Government). We utilized a quantitative method (document review) to abstract secondary data from the standardized ministry of health tuberculosis registers (HMIS 009).

We chose these health facilities as our study sites for several reasons. First, they treated >10 TB patients/per month. Second, they had a TB treatment success rate of < 75% from 1^st^ January 2019 - to 31^st^ December 2021. Third, they had both outpatient (OPD) and inpatient (IPD) clinics. Finally, they are all located within the Mukono district.

### Patient Eligibility and Data

In this study, the objective was to use machine learning techniques to identify important variables predictive of TB treatment non-adherence from a set of 42 patient demographics and clinical variables created and standardized by the ministry of health. These patient variables are collected during the TB patient treatment course. Further, these patient characteristics are recorded in a health management information systems (HMIS) register referred to as HMIS 009.

The HMIS 009 register for the period starting 1^st^ January 2019 to 31^st^ December 2021 was our main data source. We developed an electronic data capture screen using the Kobo Collect (v2021.3.4) platform. Kobo Collect is open-source software with both online and offline electronic data capture capabilities (61). The tool was pre-tested before roll out to the study. Through the tool, we aimed to abstract >1000 patient records from the drug-susceptible pulmonary TB treatment units at the study sites. These were the patients who visited the study site with a primary diagnosis of tuberculosis. They comprised both male and female patients, aged 18 years and above, who received and completed drug-susceptible pulmonary TB treatment at the study sites for the period starting 1^st^ January 2019 to 31^st^ December 2021. To meet the study objectives, and the scope and ensure data of good quality, we identified the tuberculosis patients using the inclusion criteria; 1) All patients initiated on treatment for drug-susceptible pulmonary TB, who are male or female aged > 18 years. We considered patients aged 18 years and above because younger patients (< 18 years) do not make decisions on where and when to seek health care. 2) All patients’ records with complete drug-susceptible pulmonary TB treatment outcomes are correctly filled in and meet the criteria 1. 3) All patients who received the drug-susceptible pulmonary TB treatment at the study site in the period of 1^st^ January 2019 to 31^st^ December 2021. We excluded all patients with incomplete drug-susceptible pulmonary TB treatment outcomes filled in and all patients who initiated treatment at the study sites but were later transferred out.

The data for each variable was extracted as it was presented in the registers. The dependent variable was the tuberculosis treatment outcome. This was categorized into either adherent or non-adherent. The independent variables included the client’s age, sex, category, disease classification, treatment drugs, and risk group among others.

### Data analysis

The data cleaning and basic descriptive statistics were done in STATA v15. During data cleaning, we transformed the categorical data into numerical data using numerical value labels aided by an already-defined codebook (annex 5.1). For instance, the outcome variable was abstracted as either; cured, treatment completed, treatment failure, lost to follow-up, not evaluated, or died. However, using the codebook, we transformed this into two categories with labels “1” for adherent and “2” for non-adherent patients respectively. This process was repeated for all other categorical variables within the dataset.

R studio version 2022.02.3 Build 492 and, R version 4.2.1 were utilized for modeling and building the machine learning classification algorithms. Both are freely available data analytics software. But, whereas R is an open-source, statistical, and data-centric programming language, R studio is an open-source integrated development environment (IDE), with a simple graphical user interface (UI). Furthermore, R studio provides a user-friendly point-and-click graphical user interface for the R programming language.

R provides many different algorithms for data mining and machine learning with flexible facilities for scripting experiments. We utilized the commonly used data processing libraries for data visualization, formatting, slicing, and conversion such as; “*gtsummary”*, “*tidyverse”*, and “*dplyr”*. Thereafter, we converted all the character variables into factor formats in preparation for statistical modeling. Finally, we formulated and added labels to the variables, generated a basic descriptive summary table using the “*gtsummary*” library, and, saved the prepared data ready for applying ML algorithms.

We visualized the data using basic descriptive statistics to check for incompleteness, inconsistency, and inaccuracy (errors or outliers). The analysis and modeling results were presented using tables and graphs. Lastly, the resultant clean dataset obtained after analysis was converted, exported, and stored in password-protected comma-separated values (CSV) file, before its utilization for machine learning modeling purposes.

We imputed patient demographics and clinical features records that had any missing data value. This was handled by generating a new variable called “None” and imputing all the missing values. The reason for this was that most of the missing variables were mainly laboratory tests that were not carried out and thus were not recorded. Therefore, we could not use the commonly used statistical computing techniques to estimate their respective values.

### Feature selection

Feature selection (FS) involves searching through all the attributes in the data to remove non-informative or redundant attributes whilst finding the subset of attributes that maximizes performance (62). Also, FS reduces overfitting (less opportunity to decide based on noise), improves accuracy (removing misleading variables), and reduces the training time. Due to the clinical nature of our dataset, and the research objectives, a two-stage feature selection procedure was followed. First, in building the algorithm and second, in the selection of feature importance based on the best-performing model.

In algorithm building, we applied the attribute evaluator and the filter ranker search method. In the filter ranker method, an attribute evaluator assigned a relevance score to each feature in the dataset. Thereafter, the attributes were ranked according to their relevance score in descending order. The features with a high score were selected and low-scoring features were eliminated during modeling. We tabulated the attribute selection output.

### Machine learning algorithms building

This was done in R-Studio, using the library ***“caret”***, a widely used R machine learning package. We began by splitting our dataset into two using a ratio of 80:20, 80% for training, and 20% for testing the model respectively. In the training dataset, we omitted the variables that had low scores as identified in the feature selection section above.

Next, we ran five algorithms sequentially (detailed below) based on this dataset. For every algorithm, cross-validation using a 10-fold was applied for model building with the training data set. Internally, the training dataset was split into two with 90% of the data used for training and 10% used for testing. This was repeated 10 times before the final model was built from the training dataset. The resultant model was tested using the reserved 20% testing dataset that was split from the original dataset. Using this testing dataset, we computed a confusion matrix to measure the accuracy (based on a 95% level of confidence), positive predictive value, negative predictive value, sensitivity, specificity, and kappa statistics for every machine learning model.

### Support vector machine

A support vector machine (SVM) algorithm aims to identify the maximum margin hyperplane (MMH) that creates the greatest separation between two classes (63), such that, each class has at least one support vector. Thus, to fit our SVM model, we utilized the R package - the ***kernlab*** package -, bundled in the caret package. We chose the package (kernlab) because it is developed natively in R unlike other algorithms developed in C or C++, thus allowing us to easily customize it further.

The algorithm’s (SVM) internal workings rely on vector geometry and heavy computation. But, the *kernlab* package hides this complexity from the user. Therefore, we configured the parameters required by this package and executed it. For instance, the kernel type was set to the Gaussian RBF kernel, with 231192 kernel evaluations, which has been shown to perform well for many types of data. Further, we created a custom function containing a set of custom vectors as potential cost values. We ran 12 iterations of model building, where in each iteration, a potential cost value was picked and applied from the custom vector containing the values; 0,0.01, 0.05, 0.1, 0.25, 0.5, 0.75, 1, 1.25, 1.5, 1.75, 2.

In R studio, we executed this as follows;

***model <- ksvm (target ∼ predictors, data = training_dataset, kernel = “rbfdot”, c = 0, …, 2.5)***

Where;

- **ksvm()** is a function in the kernlab package that returns an SVM object that can be used to make predictions
- **target** is the outcome in the training_dataset data frame to be modeled with.
- **predictors** specify all the features in the data frame to use for prediction.
- **data** specifies the data frame in which the target and predictors variables can be found.
- **Kernel** specifies non-linear mapping such as “*polydot*” (polynomial), “*rbfdot*” (radial basis), and linear mapping - “*vanilladot*”.
- **Cost (c)** is a number that specifies the cost of violating the constraints.

After the 12 iterations of training, each output model’s performance and accuracy were computed as the number of correct predictions divided by the total number of predictions.

### Evaluation of the developed models

For the model performance evaluation, k-fold cross-validation was used. Cross-validation is a set of methods for measuring the performance of a given predictive model on new test data sets. The rationale in cross-validation techniques is to divide the data into two sets, training sets – used to train (build) the model and the testing set (validation set) – used to test the model by computing the prediction error.

We implemented the repeated *k*-fold cross-validation method, whereby we randomly split our dataset into *k* sets. In this method, we split our data into 10-fold equal datasets. We used 9-fold (90%) datasets for training the model and the rest 1-fold (10%) of the dataset to evaluate the model performance. Thereafter, we evaluated the developed model with the test dataset (20%) to check its validity and accuracy using the unseen observations. From the results, we quantified the prediction error as the mean squared difference between the observed and the predicted outcome values.

A confusion matrix was used to measure the performance of the developed model. This is a kind of table which helps compute the performance of the classification model on a set of test data for which the true values are known.

true positives (TP) are the predicted values correctly predicted as actual positives. false positives (FP) are the predicted values incorrectly predicted as actual positives. i.e., negative values predicted as positive. Similarly, false negatives (FN) are the positive values predicted as negative. Whereas true negatives (TN) are the predicted values correctly predicted as actual negatives. The confusion matrix helps us to compute the model accuracy test using the formula below;

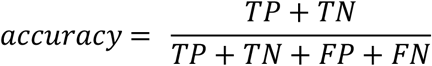

Thus, we selected the best-performing model by using a criterion of the higher the computed accuracy, the better the performance of the developed model.

Based on the confusion matrix, the model with the highest accuracy was identified. Thereafter, we computed its ***f1*** score to identify and select the critical variables. The formula; ***f1 score = 2*(Recall * Precision) / (recall + precision)*** was applied. The model with the best **f1** score, together with a sequential forward floating selection (SFFS) procedure (64) was selected for risk predictors identification.

To identify the critical variables, we first selected the best-performing model. Thereafter, we applied the Shapley additive explanations (SHAP) technique to it. SHAP is a “unified framework for interpreting machine learning models that assign each feature an importance value for a particular prediction” (65). This rationale behind the SHAP technique is that for complex models e.g. ensemble methods or deep networks, a *simpler explanation model* defined as an interpretable approximation of the original model exists.

Mathematically, SHAP applies sampling approximations to model training by assigning an importance value to each feature that represents the effect on the model prediction of including that feature. Finally, it computes the approximations of the effect of removing a given variable from the model by summing up the preceding differences for all possible subsets of all features. This is illustrated in the formula below.

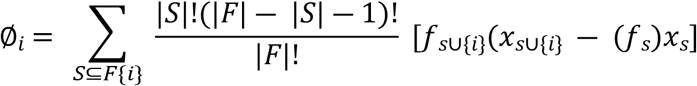

Where; ***F*** is the set of all features, *S* ⊆ *F* is the subset of all features, *f*_*s*∪{*i*}_ is the model trained with a given feature (*i*) present, while *f*_*s*_ is another model trained with the feature withheld. *x*_*s*_ represents the values of the input features in the set ***S***.

Further, the SHAP values attribute to each feature the change in the expected model prediction when conditioning on that feature (Fig 1 below). From these attributions, SHAP thus explains how to get from the base value *E*[*f(z)*] that would be predicted if we did not know any features of the current *f(x)*. This technique uses classic equations from cooperative game theory to aid in interpreting the black-box machine-learning models by computing the explanations of the model predictions.

**Fig 1.**
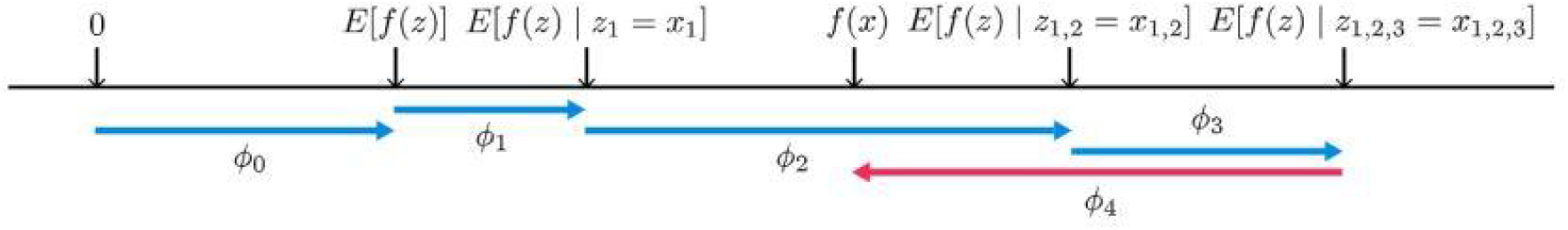
Illustrating SHAP values attribute changes of each feature in model prediction

Therefore, by applying this technique, we were able to rank our predictive features according to their sign and magnitude in response to their contribution to the outcome. Based on the feature’s contribution sign (whether positive or negative), and the magnitude, the critical predictors for either adherence or non-adherence were identified. The results were presented graphically.

## Results

In total, 1,004 tuberculosis client records were abstracted. After data cleaning, 838 records were considered for analysis and modeling. The records belonged to three (50%) government hospitals; Mukono general hospital (253), Kojja health center IV (116) and Nakifuma health center III (78) and, three (50%) private not-for-profit (PFNP) hospitals namely; Mukono COU hospital (132), Naggalama hospital (156) and Kyetume health center III (103) as shown in Fig 2 below.

**Fig 2.**
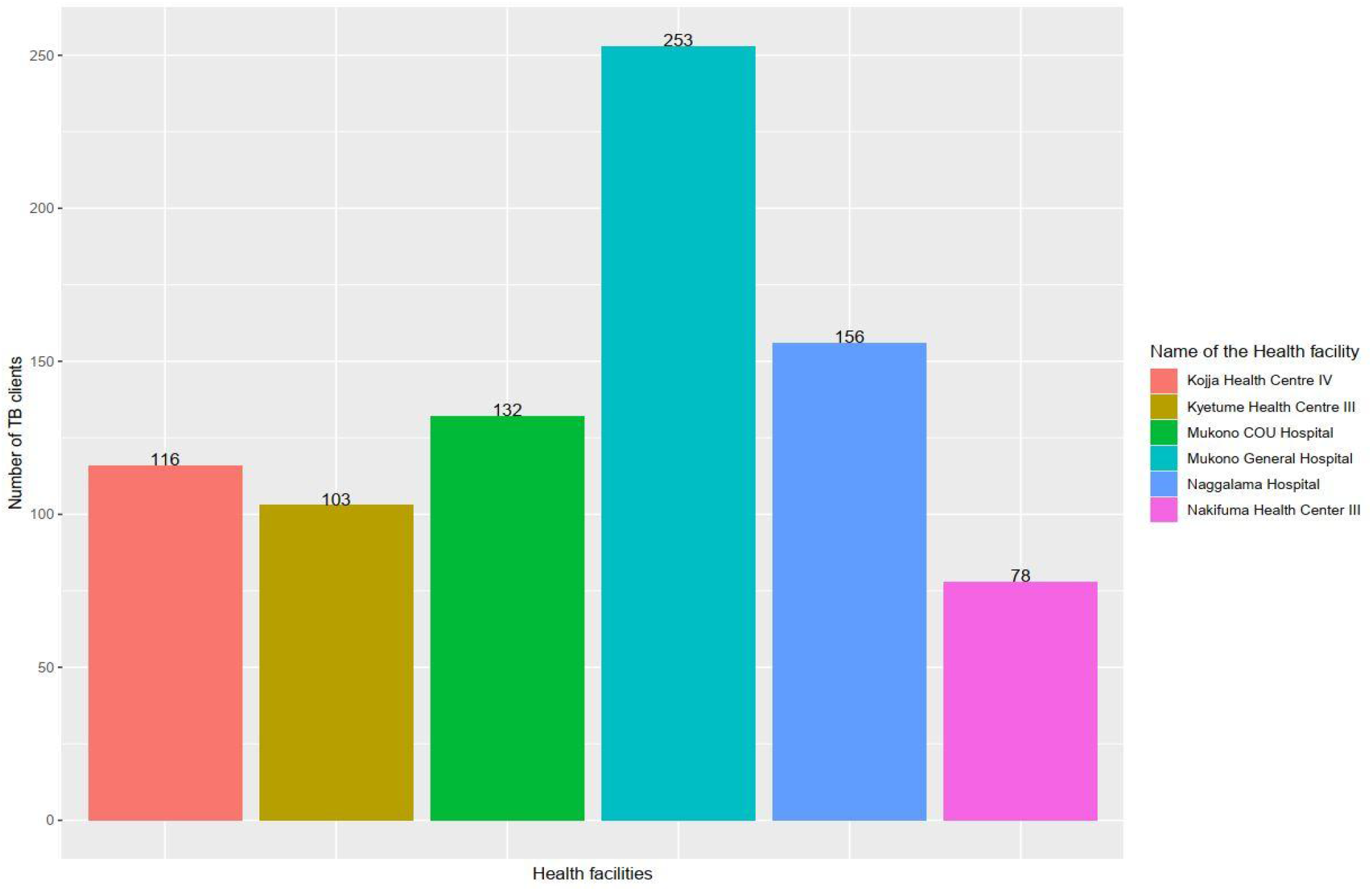
The frequency distribution of TB clients per health facility in the tuberculosis data

**Fig 3.**
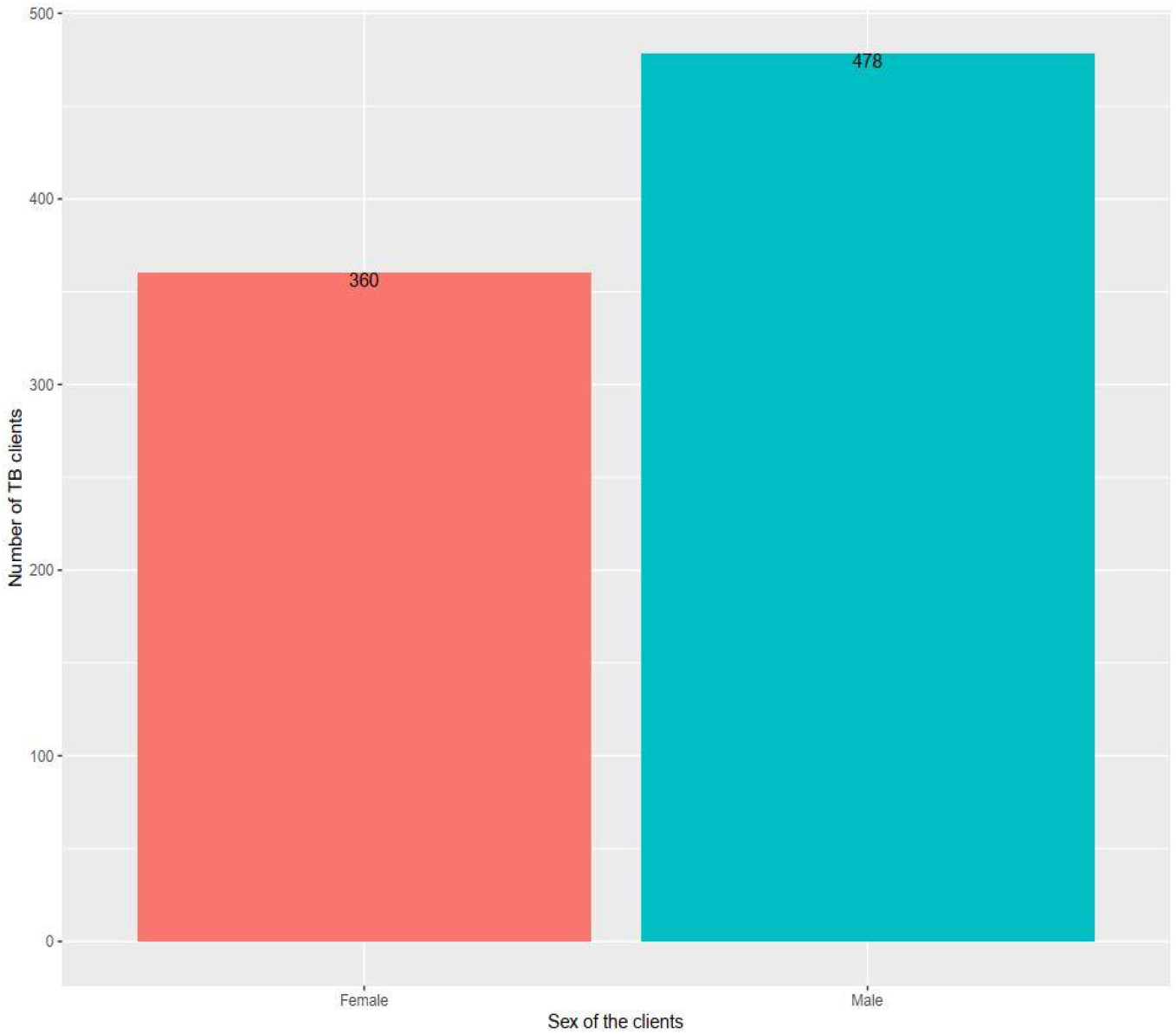
Frequency of patient sex in the tuberculosis data

The patient sex (either “male” or “female”) was balanced (57% male) as shown in figure 2. The client’s mean age was 38.3 years with a standard deviation (SD) of 13.7 years. Similarly, the client’s mean weight and standard deviation were 42.0 and 20.1 respectively. Out of all clients, 568 had contacts > 5 years (mean of 1.6 and a standard deviation of 2.2), while 340 clients had contacts <=5 years, with a mean and standard deviation of 0.7 and 1.6 respectively.

However, the categorical treatment outcome was unbalanced with 681 (81.2%) and 157 (18.8%) belonging to adherent and non-adherent classes respectively. Of all the TB tests done, the majority of the clients had GeneXpert tests (618), closely followed by smear microscopy (502) and TB LAM (39). 112 clients had no laboratory test done. A more description of the background of the study participants is presented in Table 1 below.

**Table 1:**
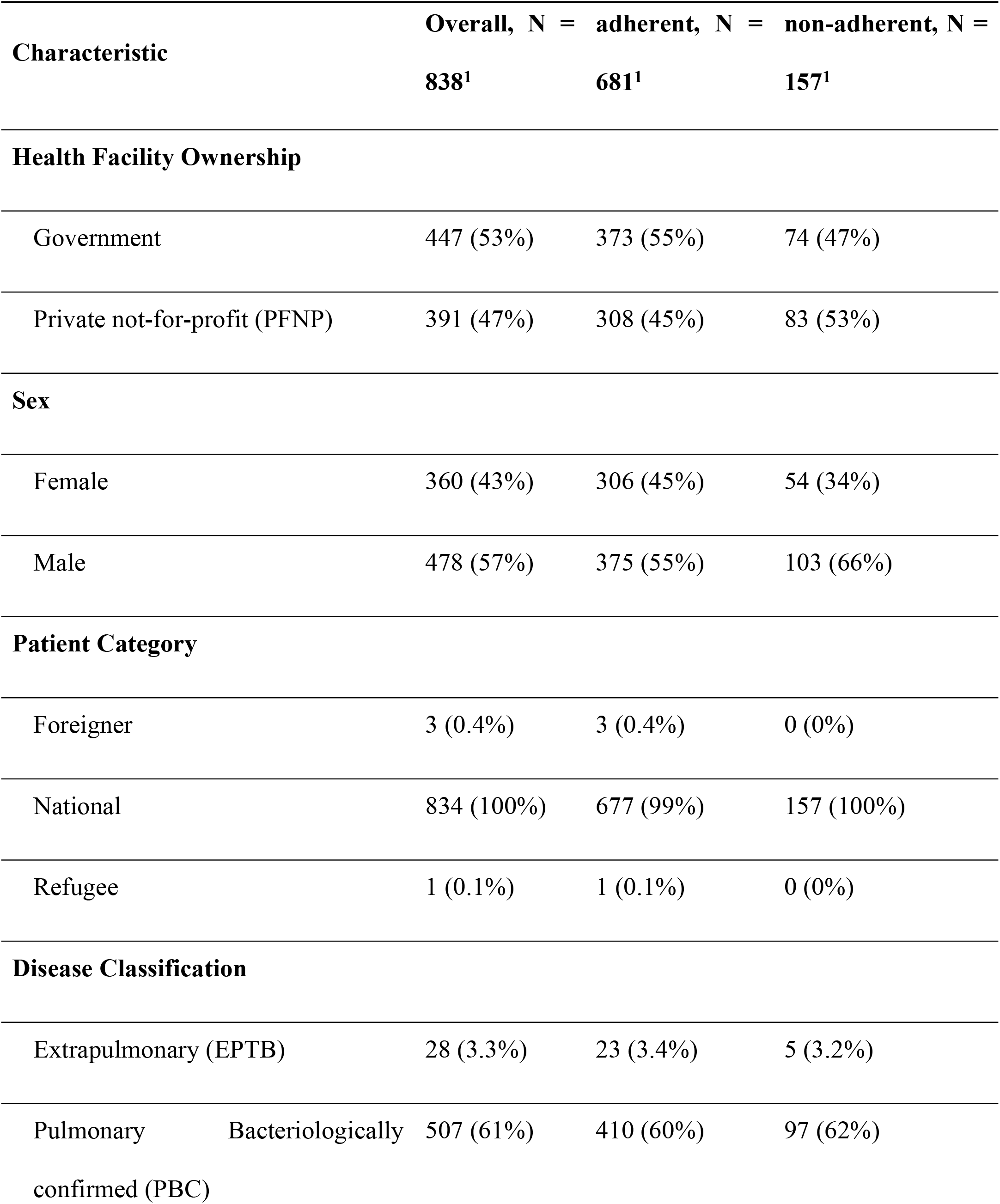
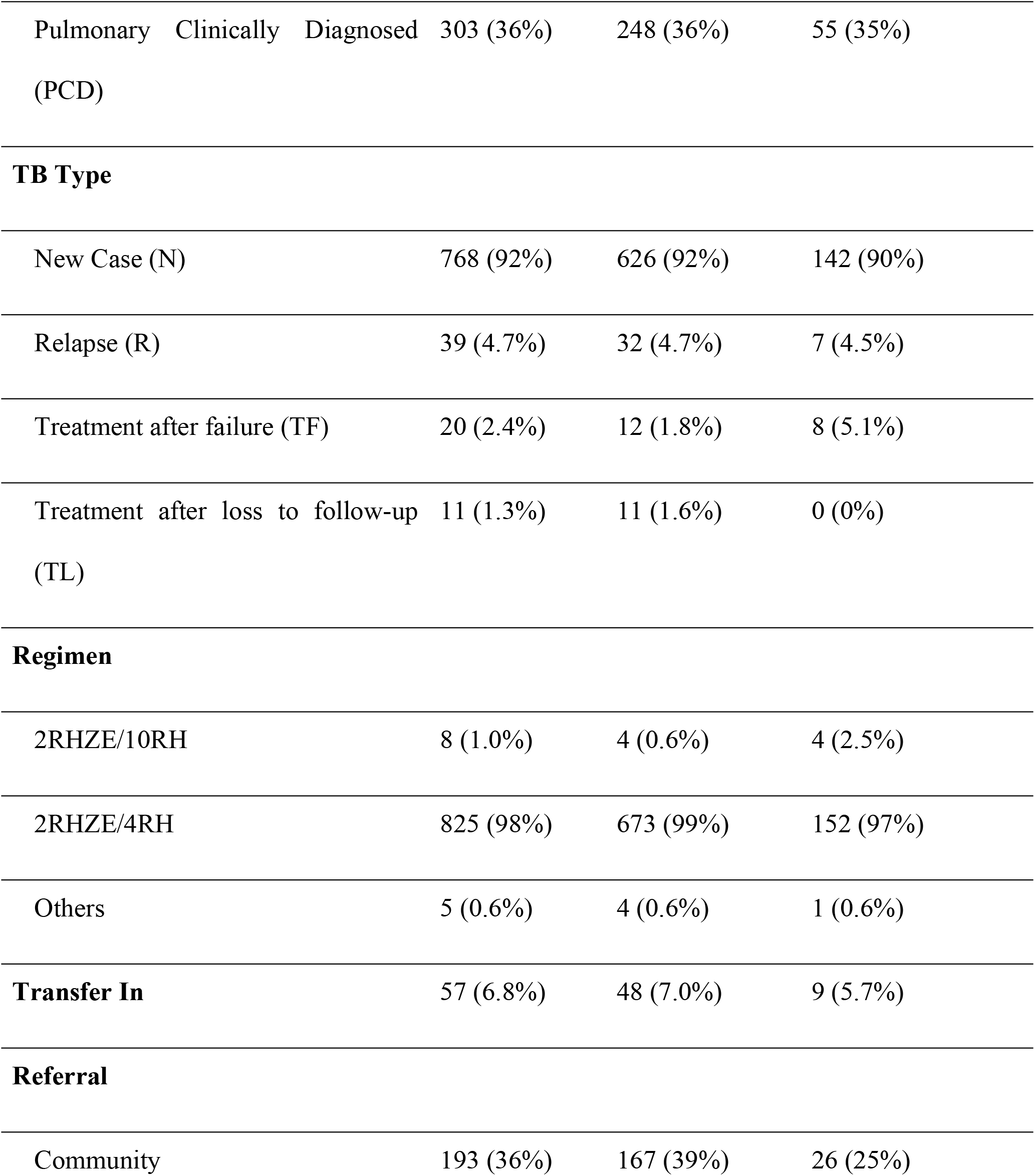
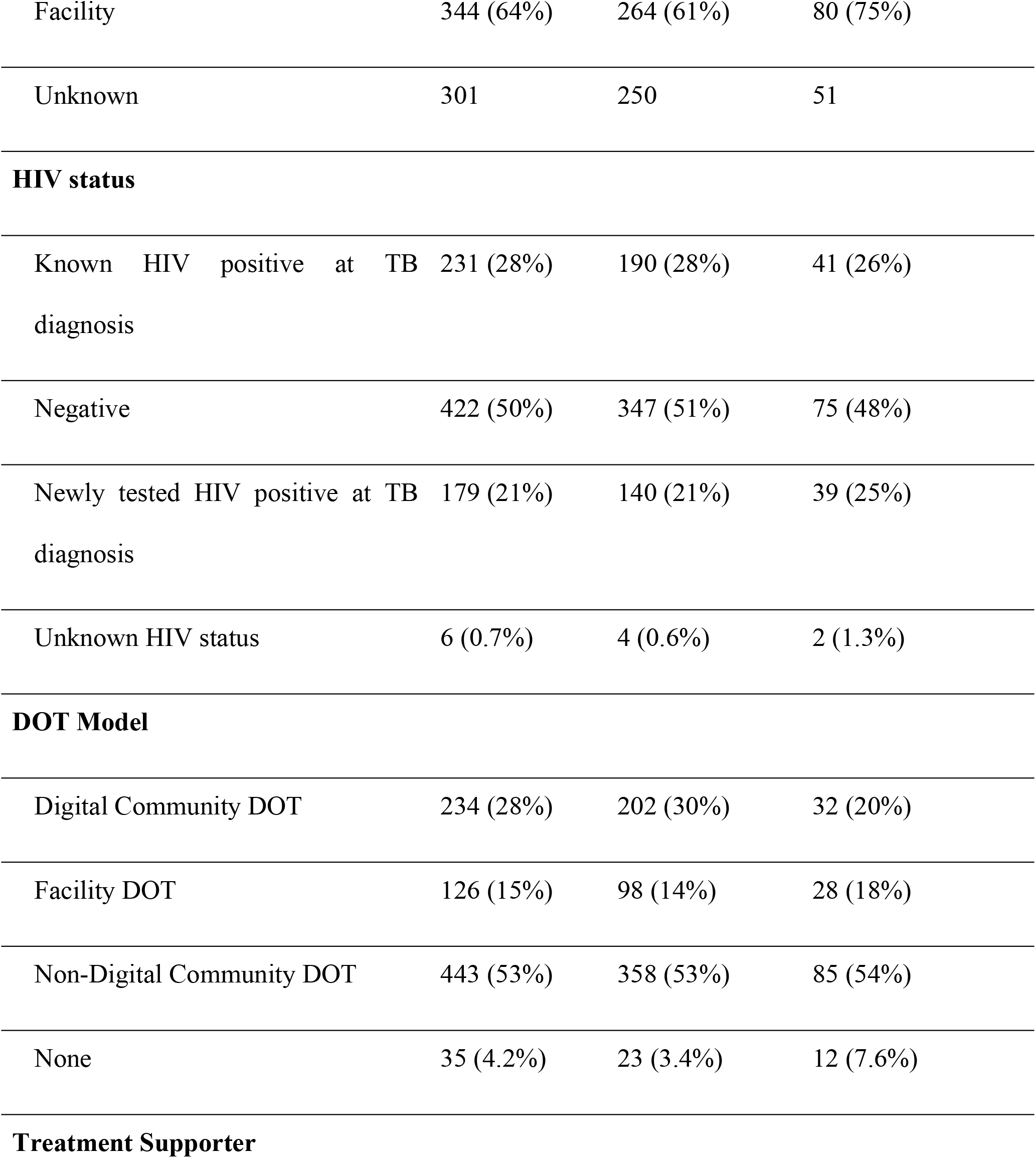
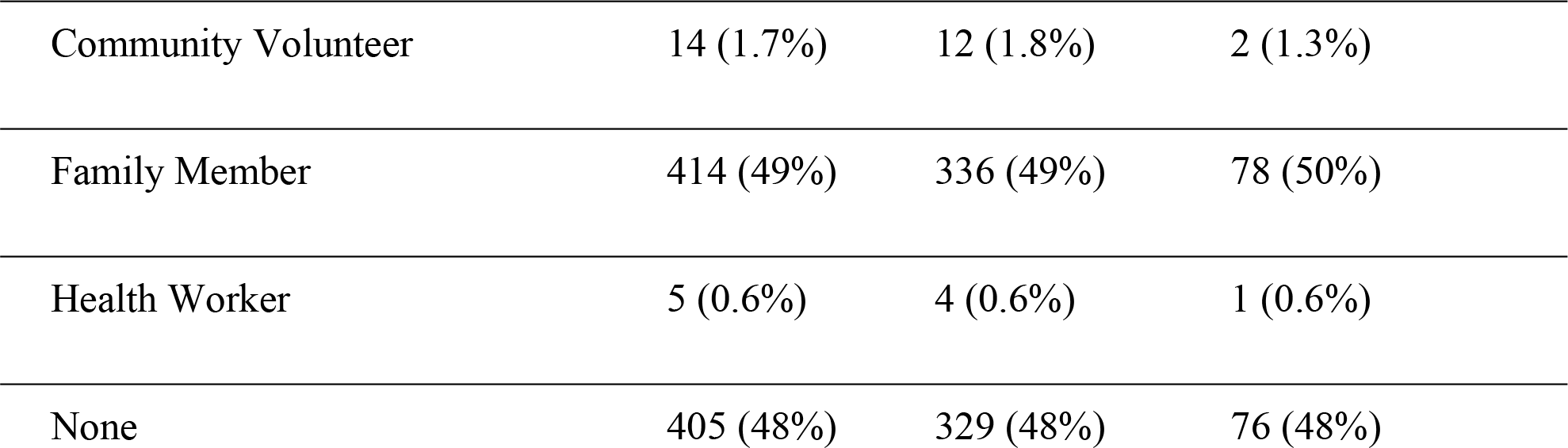
Summary statistics for key predictor and outcome variables.

We built and evaluated 5 (five) different machine-learning models. Before building and evaluating the five different machine-learning models, we conducted feature selection on all 838 instances. The resultant attribute rankings are shown in table 2 below.

**Table 2:**
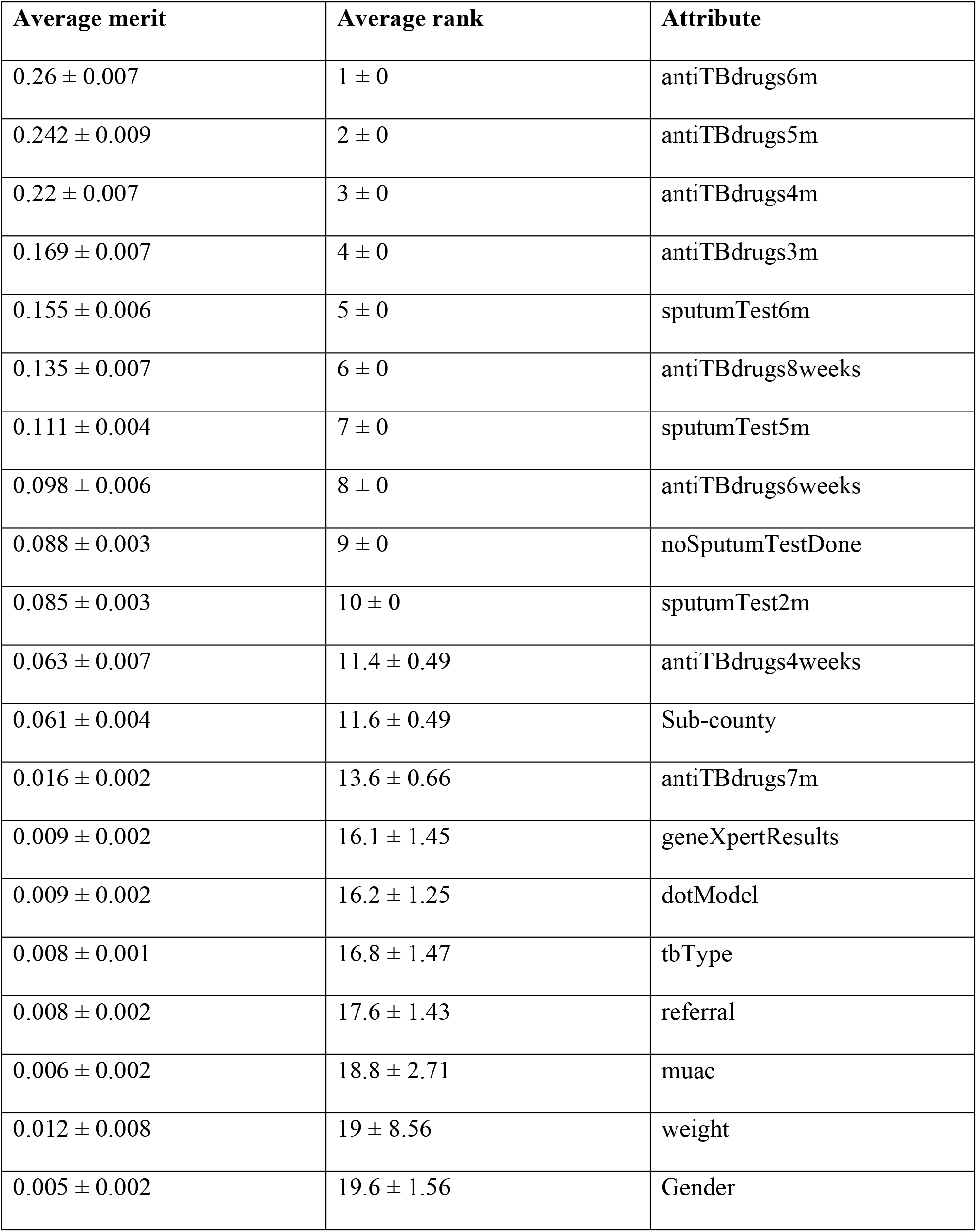
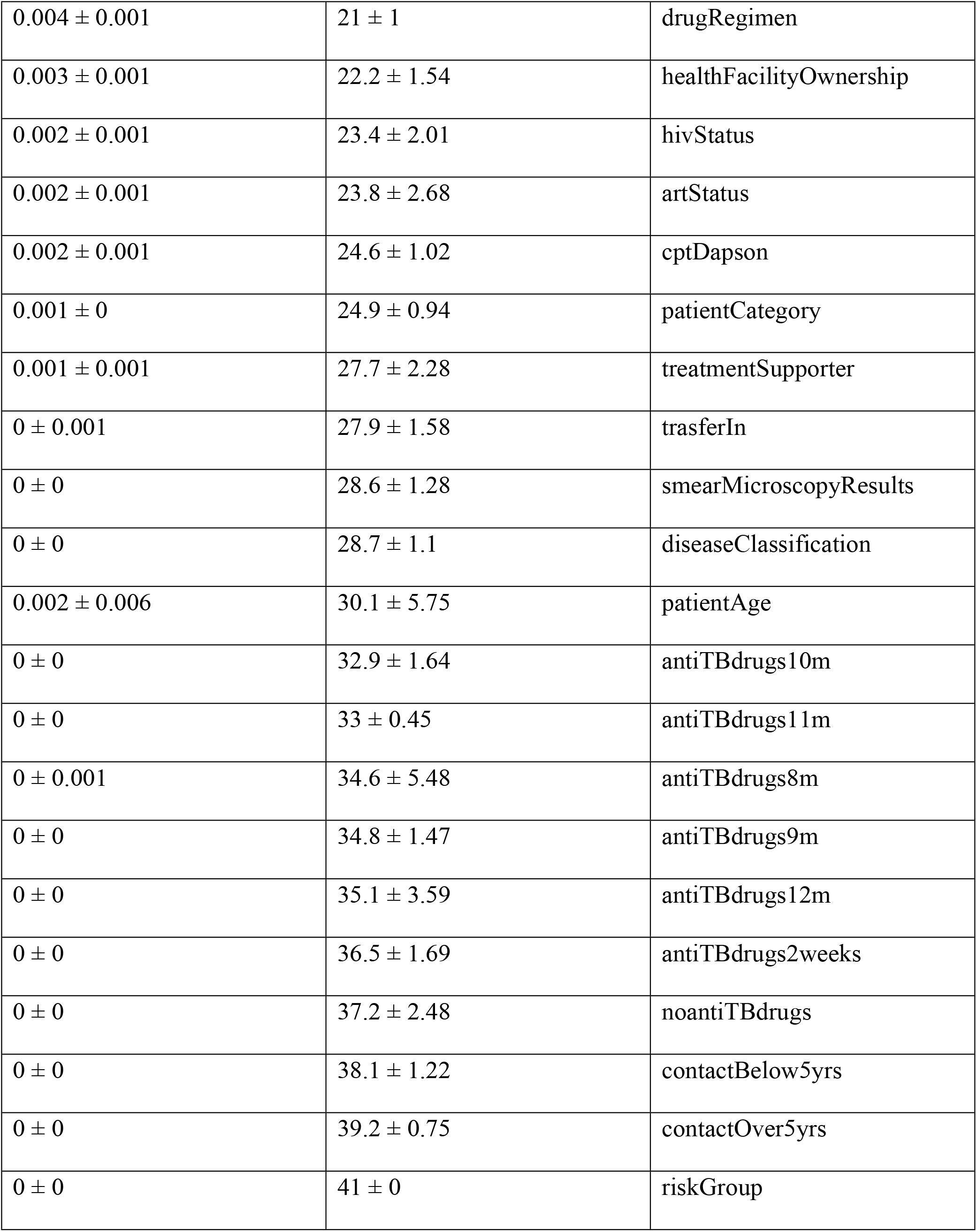
Ranking of suitable patient parameters identified through feature selection.

| Average merit | Average rank | Attribute |
| --- | --- | --- |
| $0.26 \pm 0.007$ | $1 \pm 0$ | antiTBdrugs6m |
| $0.242 \pm 0.009$ | $2 \pm 0$ | antiTBdrugs5m |
| $0.22 \pm 0.007$ | $3 \pm 0$ | antiTBdrugs4m |
| $0.169 \pm 0.007$ | $4 \pm 0$ | antiTBdrugs3m |
| $0.155 \pm 0.006$ | $5 \pm 0$ | sputumTest6m |
| $0.135 \pm 0.007$ | $6 \pm 0$ | antiTBdrugs8weeks |
| $0.111 \pm 0.004$ | $7 \pm 0$ | sputumTest5m |
| $0.098 \pm 0.006$ | $8 \pm 0$ | antiTBdrugs6weeks |
| $0.088 \pm 0.003$ | $9 \pm 0$ | noSputumTestDone |
| $0.085 \pm 0.003$ | $10 \pm 0$ | sputumTest2m |
| $0.063 \pm 0.007$ | $11.4 \pm 0.49$ | antiTBdrugs4weeks |
| $0.061 \pm 0.004$ | $11.6 \pm 0.49$ | Sub-county |
| $0.016 \pm 0.002$ | $13.6 \pm 0.66$ | antiTBdrugs7m |
| $0.009 \pm 0.002$ | $16.1 \pm 1.45$ | geneXpertResults |
| $0.009 \pm 0.002$ | $16.2 \pm 1.25$ | dotModel |
| $0.008 \pm 0.001$ | $16.8 \pm 1.47$ | tbType |
| $0.008 \pm 0.002$ | $17.6 \pm 1.43$ | referral |
| $0.006 \pm 0.002$ | $18.8 \pm 2.71$ | muac |
| $0.012 \pm 0.008$ | $19 \pm 8.56$ | weight |
| $0.005 \pm 0.002$ | $19.6 \pm 1.56$ | Gender |
| $0.004 \pm 0.001$ | $21 \pm 1$ | drugRegimen |
| $0.003 \pm 0.001$ | $22.2 \pm 1.54$ | healthFacilityOwnership |
| $0.002 \pm 0.001$ | $23.4 \pm 2.01$ | hivStatus |
| $0.002 \pm 0.001$ | $23.8 \pm 2.68$ | artStatus |
| $0.002 \pm 0.001$ | $24.6 \pm 1.02$ | cptDapson |
| $0.001 \pm 0$ | $24.9 \pm 0.94$ | patientCategory |
| $0.001 \pm 0.001$ | $27.7 \pm 2.28$ | treatmentSupporter |
| $0 \pm 0.001$ | $27.9 \pm 1.58$ | trasferIn |
| $0 \pm 0$ | $28.6 \pm 1.28$ | smearMicroscopyResults |
| $0 \pm 0$ | $28.7 \pm 1.1$ | diseaseClassification |
| $0.002 \pm 0.006$ | $30.1 \pm 5.75$ | patientAge |
| $0 \pm 0$ | $32.9 \pm 1.64$ | antiTBdrugs10m |
| $0 \pm 0$ | $33 \pm 0.45$ | antiTBdrugs11m |
| $0 \pm 0.001$ | $34.6 \pm 5.48$ | antiTBdrugs8m |
| $0 \pm 0$ | $34.8 \pm 1.47$ | antiTBdrugs9m |
| $0 \pm 0$ | $35.1 \pm 3.59$ | antiTBdrugs12m |
| $0 \pm 0$ | $36.5 \pm 1.69$ | antiTBdrugs2weeks |
| $0 \pm 0$ | $37.2 \pm 2.48$ | noantiTBdrugs |
| $0 \pm 0$ | $38.1 \pm 1.22$ | contactBelow5yrs |
| $0 \pm 0$ | $39.2 \pm 0.75$ | contactOver5yrs |
| $0 \pm 0$ | $41 \pm 0$ | riskGroup |

**Table 3:** Developed models evaluation results.

| No. | Prediction algorithm | Evaluation parameters |  |  |  |  |
| --- | --- | --- | --- | --- | --- | --- |
|  |  | Precision | Recall | Accuracy (%) | F1 score | ROC |
| 1. | Logistic regression | 0.8850 | 0.8830 | 88.30 | 0.884 | 0.896 |
| 2. | Support vector Machine<br>(SVM) | 0.9160 | 0.9130 | 91.28 | 0.914 | 0.870 |
| 3. | Random Forest (RF) | 0.8980 | 0.9000 | 89.97 | 0.665 | 0.929 |
| 4. | AdaBoost | 0.9130 | 0.9110 | 91.05 | 0.911 | 0.920 |
| 5. | Artificial Neural Networks<br>(ANN) | 0.8790 | 0.8830 | 88.30 | 0.881 | 0.902 |

After the feature selection, we developed five models based on the resultant dataset. Thereafter, we computed the metrics (precision, recall, accuracy, receiver operating curve (ROC), and, F1 score) of these developed models. Table 2 below shows the results of these metrics obtained per model.

From the above findings, the support vector machine (SVM) had the best accuracy of 91.28%, closely followed by AdaBoost (91.05%), and Random Forest (89.97%). Both Artificial neural networks (ANN) and Logistic regression had equal accuracy of 88.30%. However, Random forest had the lowest F1 score of 0.665 compared to other algorithms that scored greater than 0.800.

In our SVM modeling, we tuned the classifier to get better performance with different values of C (cost) per kernel function. With the linear kernel function, we obtained the best model accuracy of 91.28% with a cost parameter of c = 0.1, among the twelve models evaluated as shown in Fig 4 below. Further exploration revealed that the model accuracy decreased with increased cost.

**Fig 4.**
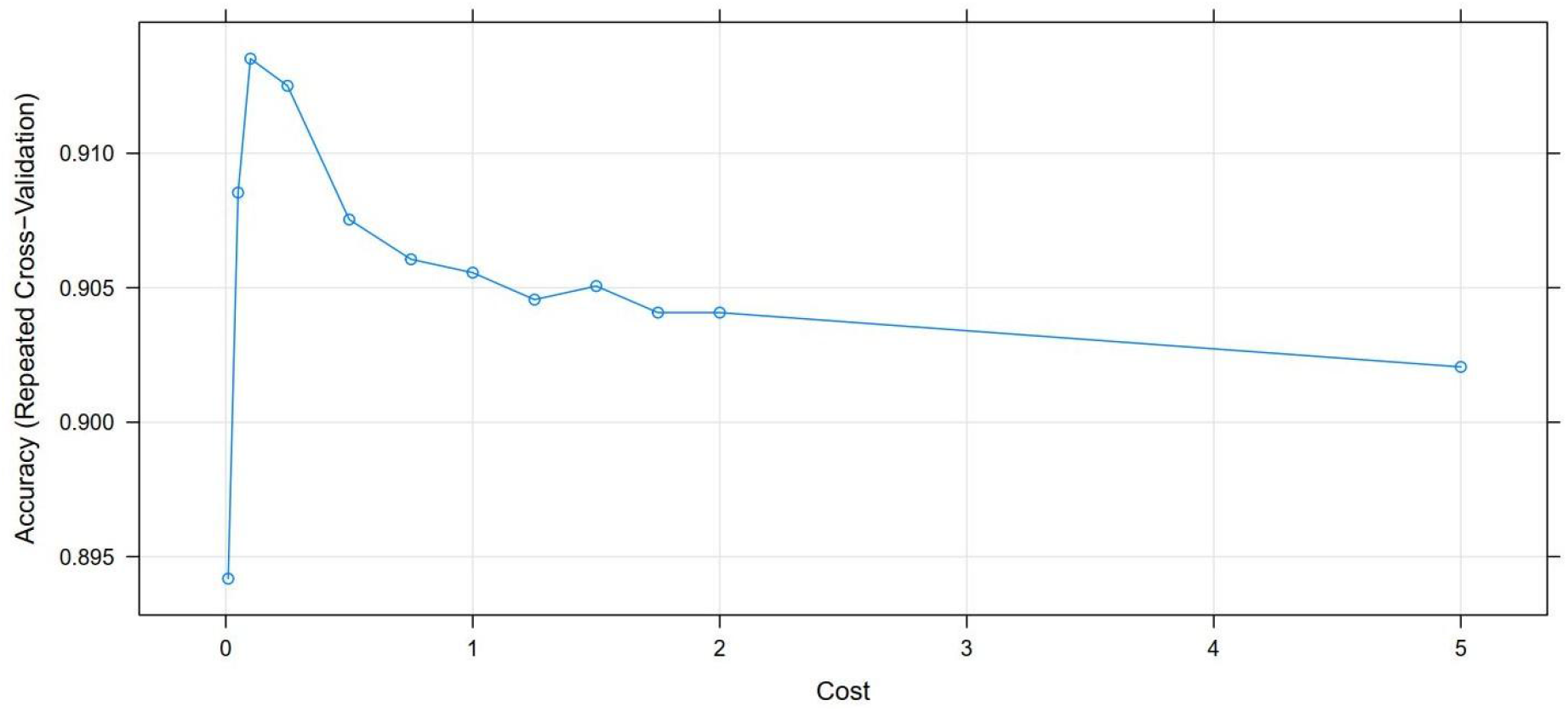
Accuracy versus the cost of the support vector machine model

Out of the five, the support vector machine (SVM) model had the best performance. Thus, we applied the Shapley technique (SHAP) to this model. This was to identify the important variables that could estimate the tuberculosis treatment non-adherence. The SHAP technique ranked all the variables used for modeling based on their contribution toward the tuberculosis treatment outcome. These are shown in Fig 5 below;

**Fig 5:**
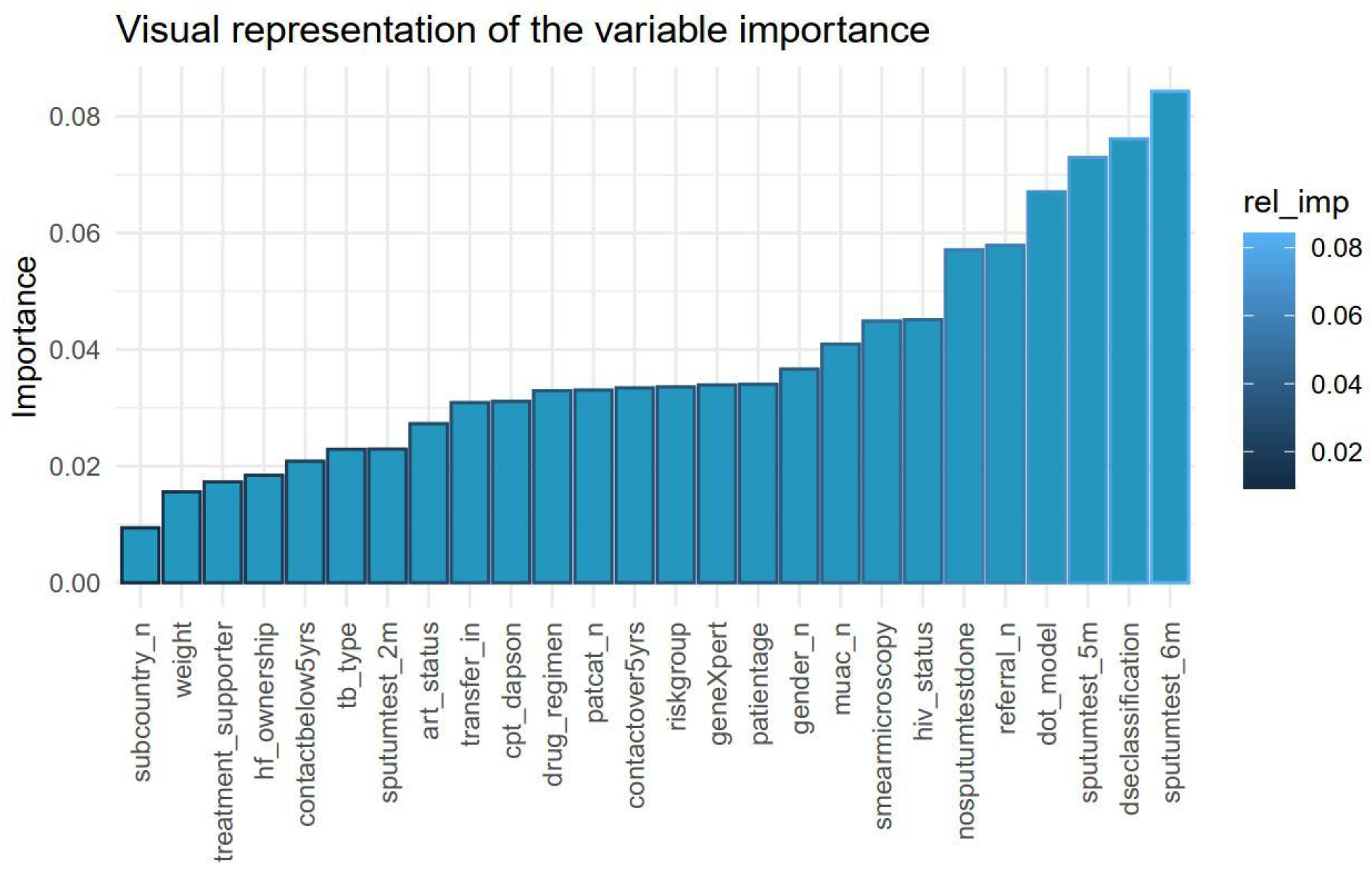
Visualization of the independent variables’ importance using the Shapley technique

On further exploration, the Shapley technique identified predictors’ importance based on their magnitude and sign (positive and negative) in relation to their contribution. The positive predictors were associated with adhering, while negative predictors are associated with non-adherence (Fig 6).

**Figure 6:**
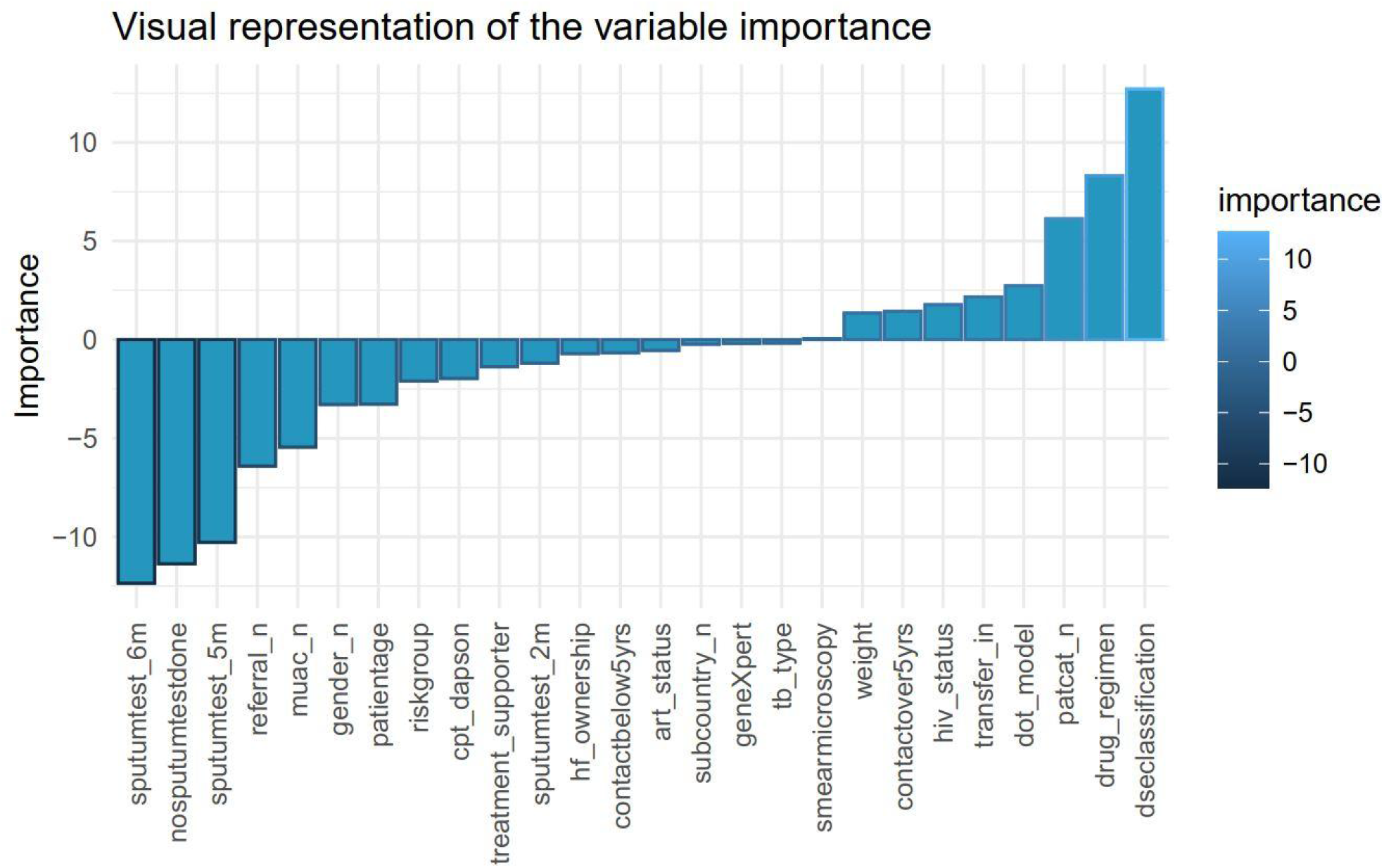
Visual representation of the positive and negative predictors

From the graphical representation above, disease classification, drug regimen, patient category, DOT model, transfer-in, HIV status, contacts over 5 years, weight, and smear microscopy results were identified to contribute positively towards treatment adherence.

In contrast, TB type, GeneXpert results, sub-country, ART status, contacts below 5 years, health facility ownership, sputum test results at 2 months, treatment supporter, CPT Dapson status, risk group, patient age, gender, middle and upper arm circumference (MUAC), referral, positive sputum test at 5 months and 6 months were indicative of treatment non-adherence.

## Discussion

Our study focused on developing and evaluating different machine learning algorithms to explore the risk factors predictive of treatment non-adherence. Thus, after training the models, we evaluated them for performance and accuracy. The metrics applied were; precision (false positive), recall (false negative), F1 score, accuracy, and receiver operating curve (ROC). However, due to the clinical nature of our study, we were interested in a model with high recall. That is, we wanted a model capable of identifying the highest number of patients who would not adhere to treatment, even if that meant having some false positives (patients wrongly identified as not adherent yet they are).

In a real clinical setting, false positives may lead to additional unnecessary clinical examinations, and extra laboratory tests and even frighten some patients. However, the public health benefits outweigh this setback. This is because the additional tests could help identify and take care of other unaware sick people thus reducing possible community spread, transmission, and possible death.

Our support vector machine (SVM) model yielded a high accuracy (91.28%) and high recall (0.9130) compared to other ML algorithms (table 5 in the results section) in classifying treatment non-adherence. However, the high recall attained by the SVM model was expected. This is due to the internal workings of the SVM algorithm that aims to identify a hyperplane that separates the training data, by maximizing the gap between these hyperplanes so that in case of any new data, it’s mapped with a maximum gap separating the classes.

Indeed, these characteristics exhibited by SVM have led to its wide adoption and utilization in several studies (32,52,54,66) that aimed to predict treatment failure and risk factors in tuberculosis and other illnesses. In all these studies, SVM algorithms were reported to have attained high accuracy and recall like ours. Thus, our findings of SVM as the best performer further confirm its robustness as a classification algorithm.

Noteworthy, all the five machine learning algorithms investigated were able to discriminate between the outcome class with high precision (> 0.87) considering all the 41 study features.

These results indicate that classification machine learning models can be used to map the different predictors to their respective classes. Likewise, these capabilities have been reported in similar rather challenging scenarios like predicting viral failure (53), tweets classification for disease surveillance (59), identification of HIV predictors for screening (60), and dermatology conditions (67). Therefore, this demonstrates the potential and applicability of machine learning algorithms to provide insights in scenarios where human decision-making would be limited.

A deeper exploration of the best-performing SVM model identified some variables predictive of tuberculosis treatment non-adherence. We relied on the SHAP technique’s measurement for each independent variable contribution to the treatment outcome for the identification of these variables.

From our findings, TB type, GeneXpert results, sub-country, ART status, contacts below 5 years, health facility ownership, sputum test results at 2 months, treatment supporter, CPT Dapson status, risk group, patient age, gender, MUAC, referral, positive sputum test at 5 months and 6 months were predictive of treatment non-adherence. These results are like other studies conducted in lower- and middle-income countries exploring the patient characteristics associated with treatment non-adherence.

Patient age and gender had equal importance in contributing to treatment non-adherence. This is possible because gender-based roles and responsibilities increase with the increase in age (15,25). As men mature, they start engaging in income-generating activities that are not only demanding but also time-consuming. In turn, this may hinder men from going to health facilities to pick up their drug refills or even taking medicines as prescribed. Similarly, women also take up responsibilities like childbearing, and household chores that equally may hinder them from going for drug refills.

Various studies have reported on the significance of patient risk groups and residency as key determinants in treatment adherence (11,50,68,69). In our modeling, we equally found the two characteristics to be predictive of whether a patient will adhere to treatment or not. Further, we found out that some patients who were not residents of our study site catchment areas attended the health facilities located within Mukono. This could be because of the stigma associated with tuberculosis as was discussed by (69). In a similar scenario for covid-19 treatment seeking, a study by Muttamba et. al.. (70) also cited stigma as a major hindrance. However, we noted that the grouping of risk groups as others could have masked other types of patient risk categories that could be on tuberculosis treatment and probably not adhering.

Whether a patient had a treatment supporter or not was found to be predictive of treatment non-adherence. Previous findings identified the important role played by treatment supporters in supporting chronic patients to adhere to treatment (10,13,31). With the patient at liberty to choose between either a family member, community health worker or health worker as a supporter, the presence or lack of one thereof greatly impedes the treatment taking.

Comorbidity of tuberculosis and HIV/AIDs is another important factor that contributes to treatment non-adherence. From our findings, both ART status and CPT Dapson status were among the factors identified by our model. Other researchers’ findings (8,27,71) showed that the drug burden for these chronic and infectious diseases can overwhelm the patient who either decides to take drugs for one of the diseases and not take the others based on either drug reactions or side effects.

MUAC measurement, an indicator of patient nutrition status was also found to be predictive of non-adherence. This concurred with two different studies (72,73) both conducted in Uganda, among tuberculosis patients, that found that undernutrition leads to unfavorable treatment outcomes. This finding also resonated with the fact that tuberculosis drugs are administered according to patient weight at diagnosis. However, upon treatment initiation, tuberculosis drugs have been shown to affect the patient’s appetite.

## Conclusion

In TB management, treatment non-adherence remains prevalent. Though the existing treatment adherence strategies advocate for active supervision and support for all TB patients, this may not be possible due to the cost implications and limited resources. However, our study findings indicate that supervised machine learning models are capable of discriminating between adherent and non-adherent clients, with high accuracy and high recall. Thus, machine learning can potentially be used as a regular tool to explore individual risk factors for tuberculosis treatment non-adherence.

Furthermore, we built all five machine learning models using the data collected routinely in a typical tuberculosis clinic in LMIC. On further experimentation of these datasets, using the feature extraction technique, we were able to identify some predictors of treatment non-adherence. Therefore, classification machine learning algorithms mainly; logistic regression, support vector machine (SVM), random forest (RF), AdaBoost, and artificial neural networks (ANN), can be built from the readily available health facility registers data.

Finally, whereas we front machine learning as an alternative advanced technique for finding hidden patterns in data, it comes at a cost. This cost is incurred in skillsets requirement, computing infrastructure, data collection, cleaning, labeling, wrangling, and modeling processes. Thus, this not only calls for preparations to incur the mentioned costs but also to invest time to experiment with different machine learning algorithms. This is to ensure that persons with the right skill set, computing infrastructure, and data of high quality can build machine learning models that can generate insights, to guide in both decision-making and policy formulations to better healthcare in our communities.

### Limitations

Despite the study findings, we encountered some limitations. First, the use of TB registers as a data source may not fully describe/capture the entire non-adherence process. This is partly because these registers leave out some social-environmental, social-demographic factors like patient occupation, marital status, number of children, and distance from the health facilities that have been shown by other qualitative studies to be associated with non-adherence. However, we successfully identified some important factors that could guide non-adherence screening despite the lack of environmental and social-demographic factors.

Likewise, the heterogeneous classification of “adherent” and “non-adherent” patients further complicates the interpretability of our results. For instance, we classified unpredictable events like the death of the client as non-adherent. However, this event could have occurred due to other factors not necessarily related to TB treatment. Thus, this limits the wider application in such scenarios. The poor data quality limited our deeper modeling and analysis. For instance, the risk group “others” instead of the actual category hindered us from modeling the actual risk that could be attributed to any given risk group categories other than the commonly known groups. Similarly, client laboratory tests varied according to health facilities with some not carrying out any. The lack of uniform laboratory tests and missing laboratory tests done in the health facilities hindered our comparison metrics.

### Recommendations & future work

We suggest that future work should collect more data from a bigger population, to enrich the dataset for modeling treatment non-adherence. More data could be collected qualitatively through administering key informant interviews to both the health workers and the patients. This qualitative data, though needing prior data transformations will help in better understanding the complex role of human behavior in treatment non-adherence. Further, it will greatly inform and supplement our findings to get a complete picture of the risk factors for tuberculosis treatment non-adherence.

Second, for the tuberculosis management programs, we recommend an additional field in the HMIS 009 registers to capture the actual risk group of the client instead of employing the “others” option. Further, identified tuberculosis clients with several TB contacts > 5 years exposed to TB through household contact should be prioritized for screening so they can be started on treatment to prevent infection or progression of the disease.

Third, to the hospital management, frequent data quality assessments should be performed to address issues in missing data variable entries in the registers. Additionally, this will ensure that the management can address some issues raised by the data staff like patients missing contact numbers or modalities of treatment support. Also, the health workers in the tuberculosis treatment units should be trained on the importance of health communication and awareness so that they can relay the same to their clients.

Fourth, we recommend the MoH and the health facility in-charges adopt information, communication, and technology (ICT) for tuberculosis management to ease the workload of filling the manual paper-based registers. This will not only improve data quality due to readily available checks in the electronic system but also enable real-time monitoring and evaluation of tuberculosis programs both in the health facility and country-wide.

Finally, we implore future researchers to carry out digital health innovations feasibility, and acceptability tests within the health facilities. Initial assessments can be targeted to the health workers to validate and identify the bottlenecks to scaling digital health innovations and clinical decision support systems like our machine learning models in both government and private-not-for-profit health facilities.

### Ethical approval

Ethical approval was sought and granted (approval protocol number 047) by the Institutional Review Board (IRB) at Makerere University School of Public Health to conduct the research. In addition to the approval, we were granted permission by the Mukono district health officer (DHO) to carry out the study in the six (6) study sites. Further permission from the health facilities in-charge and the director at Mukono General Hospital and Mukono COU Hospital were sought and granted.

## Data Availability

All relevant data are within the manuscript and its Supporting Information files.

## Acknowledgments

To Mukono District Health Officer (DHO) who permitted us to conduct and collect data for this study in his district. Special gratitude to the management of Kyetume Health Centre III, Mukono General Hospital, Naggalama Hospital (PNFP), Mukono COU Hospital, Kojja Health Centre IV, and Nakifuma Health Centre III.

Ivan Mwesigwa, Mukono district biostatistician, and the respective health facilities data officers for the support rendered during the data abstraction process.

To Dr. Vincent Kiberu, Dr. Alice Mugisha, and Dr. Ronald Muhumuza Kananura who provided immeasurable insights and advice during the entire study period.

## Notes

### Competing Interest Statement

The authors have declared no competing interest.

### Funding Statement

The author(s) received no specific funding for this work

### Author Declarations

Makerere University School of Public Health Institutional Review Board

